# Race Corrections in Clinical Algorithms Can Help Correct for Racial Disparities in Data Quality

**DOI:** 10.1101/2023.03.31.23287926

**Authors:** Anna Zink, Ziad Obermeyer, Emma Pierson

## Abstract

Despite ethical and historical arguments for removing race corrections from clinical algorithms, the consequences of removal remain unclear. An important and underdiscussed consideration in this debate is the fact that medical data quality frequently varies across race groups. For example, family history of cancer is an essential predictor in cancer risk prediction algorithms but is less reliably documented for Black patients and may therefore be less predictive of cancer outcomes. We assessed whether race corrections could allow risk prediction models to capture varying data quality by race, focusing on colorectal cancer risk prediction. Using data from the Southern Community Cohort Study, we analyzed 77,836 adults with no history of colorectal cancer at baseline. We assessed whether the predictive relationship between self-reported family history of colorectal cancer and 10-year colorectal cancer risk differed by race. We then compared two cancer risk prediction algorithms -- a race-blind algorithm which included standard colorectal cancer risk factors but not race, and a race-corrected algorithm which additionally included race. Family history predicted 10-year colorectal cancer risk among White patients (OR: 1.74, 95% CI 1.25-2.38), but not Black patients (OR: 0.98, 95% CI 0.72-1.29). Relative to the race-blind algorithm, the race-corrected algorithm improved predictive performance, as measured by goodness of fit in a likelihood ratio test (p-value <0.001) and AUROC among Black patients (0.611 versus 0.608, p-value 0.006). Because the race-blind algorithm underpredicted risk for Black participants, the race-corrected algorithm increased the fraction of Black participants among the predicted high-risk group, potentially increasing access to screening. Race corrections can allow risk prediction algorithms to model varying data quality by race group, which frequently occurs in clinical settings.

## INTRODUCTION

The medical community is locked in a consequential debate over when clinical algorithms should use *race corrections,* which include patient race as an input to the algorithm. This debate has immediate implications for patient care across domains including cardiology, pulmonology, nephrology, urology, obstetrics, oncology, and endocrinology, which have historically relied on algorithms containing race corrections.^1^ For example, some health systems have recently removed race as an input when estimating glomerular filtration rate^2^, a measure of kidney function which guides diagnosis of kidney disease, drug recommendations, and kidney transplants.^3^ The reconsideration of race corrections is vital and long overdue: some race corrections rely on dubious data, exacerbate health disparities, or flow from false beliefs about race as a biological variable, when it is in fact a social construct.^1^ At the same time, the consequences of removing race corrections from clinical algorithms remain unclear, and researchers have called for caution and careful study before doing so.^4–6^

Here, we study an important and largely undiscussed consideration in the race corrections debate: varying data quality across race groups. Differences in medical data quality by race group, and their consequences for health equity, occur frequently and have been documented in diverse domains.^7^ When input variables to clinical algorithms are less reliably recorded for some race groups, these variables will tend to have less predictive power for those race groups.^8–10^ Race-blind models may fail to capture this, relying too heavily on the unreliable input features for race groups with worse data quality; in contrast, race corrections can allow predictive algorithms to capture data quality which varies by race group.

One example of this phenomenon is family health history data. Family history of cancer is a known risk factor for many cancers.^11^ But *recorded* family history data, often collected during medical interviews with a clinician, varies in quality across race groups. A number of studies have found that family history of cancer is better known and documented in White patients.^12–15^ These racial disparities in data quality mean that the predictive value of recorded family history could vary across race groups. In particular, the absence of recorded family history may be less reassuring in non-White patients, who may be incorrectly recorded as having no family history either because the clinician does not ask, or the patient does not know. A race-blind risk prediction would fail to account for this, producing inappropriately low predicted risks for non-White patients without recorded family history; in contrast, a race correction could capture how the prognostic value of recorded family history varies by race and improve prediction. We tested this hypothesis on a colorectal cancer risk score, where family history of colorectal cancer is an important risk factor and typically results in earlier and/or more frequent screening.^16^

## METHODS

### Data

Our data come from the Southern Community Cohort Study (SCCS) established in 2001 to study cancer disparities as well as other health conditions in the southeastern U.S.^17^ SCCS enrollment began in 2002 and continued for eight years (until 2009). Patients were primarily recruited from community health centers in the following twelve states: Alabama, Arkansas, Florida, Georgia, Kentucky, Louisiana, Mississippi, North Carolina, South Carolina, Tennessee, Virginia, and West Virginia. Data were collected from surveys administered at the time of enrollment and several follow-up periods. Data from the baseline survey were collected either through a self-administered survey or an in-person computer-assisted interview. Follow-up surveys were done by telephone or self-administered: approximately 68% of patients completed the follow-up surveys. State cancer registry data were linked to patients when possible.

The primary outcome was whether the participant developed colorectal cancer in the ten years following enrollment. This variable was measured using the follow-up survey, cancer registry data and National Death Index reports of malignant neoplasms of the colon, rectum, and anus. We included all recorded cases of colorectal cancer from any of these three sources.

Family history of cancer was collected for patients’ birth mother, birth father, full sisters, and full brothers. For each family member, respondents could select “yes”, “no”, or “don’t know” for whether the person had cancer. Respondents who indicated that any of these family members had cancer then selected which type of cancer they had. We defined a participant as having a known family history of colorectal cancer if they indicated that one of their family members had colorectal cancer, consistent with previous work.^18^ For the main analysis, we compared participants with a known family history of cancer to participants who did not have a known family history of cancer (grouping the “don’t know” and “no family history” respondents together in the latter category). In a sensitivity analysis, we considered the effects of two alternate ways of coding family history: 1) analyzing family history as a 3-level categorical variable with “don’t know” as a separate category and 2) grouping the “don’t know” group with the “yes” group as opposed to with the “no” group.

All covariates were measured using data collected in the baseline survey. We defined race groups based on the participants’ description of their race or ethnicity at baseline. Participants had six options to choose from (White, Black/African-American, Hispanic/Latino, Asian or Pacific Islander, American Indian or Alaska Native, and Other racial or ethnic group) and could mark all that apply. We defined Black participants as any participants who described themselves as Black/African-American. We defined White participants as any participants who described themselves as White only. More than 95% of the sample identified as either Black or White only, so we only included participants in these two groups for the analysis, following previous work.^19^

### Analysis

We examined the prognostic value of family history by race group by plotting 10-year colorectal cancer rates by age, since age is an important risk factor for colorectal cancer that affects screening recommendations.^20^ We also ran separate logistic regressions for Black versus White participants in which we predicted 10-year colorectal cancer incidence given age and family history and reported the odds ratio on the family history coefficient for each regression with 95% confidence intervals estimated using profile likelihood methods.

Then, we created two screening algorithms that modeled 10-year colorectal cancer risk as a function of age, sex, family history, screening history, and lifestyle habits based on the set of controls used in the NIH Colorectal Cancer tool: participant age at the time of enrollment, an indicator for female, BMI greater than 30, ever had a sigmoidoscopy, ever had a colonoscopy, ever had polyps, the age that the polyp was identified if ever, smoking status (current, former, never), drinking status (<=1 drink per day, >1 drink per day), whether they took NSAIDs or Aspirin regularly, whether they did any vigorous activity, and whether they ate vegetables each day.^21^ One algorithm was race-blind (i.e., did not include race as a predictive feature), whereas the race-corrected algorithm added an indicator for whether the participant was Black both as a main effect and as an interaction with family history, in addition to the set of controls used by the NIH risk tool.^21^

We compared predictive performance of the race-blind and race-corrected algorithms using two measures. First, we performed a likelihood ratio test to compare goodness of fit in the race-corrected versus race-blind algorithm. Second, we compared the two algorithms in terms of overall and race-specific Area Under the Receiving Operating Characteristic (AUROC), a standard measure of predictive performance, on a holdout test set comprising 50% of the dataset.^22^ We tested for statistically significant improvements in AUROC using DeLong’s algorithm.^23^

To assess how the race-corrected algorithm might impact colorectal cancer screening decisions, we compared the assignment of patients to high-risk strata under the race-blind and race-corrected algorithms, since patients assigned to high-risk strata are more likely to be screened. We defined predicted high-risk participants as those in the top k% percentile of predicted risk (where k = 50, 25, 10, 5, and 1), and looked at the share of Black participants among the predicted high-risk group. Uncertainty estimates were calculated by bootstrapping the test set, and reporting confidence intervals across 5,000 bootstrap iterations.

We also performed a set of checks to ensure that reported family history remained more predictive for white patients under different outcome definitions, model choices, and definitions of family history.

First, we checked that the interaction term between family history and race remained significant under two additional outcome definitions to check that the results were not sensitive to the source of reported data: (1) colorectal cancer cases reported in the follow-up surveys, and (2) colorectal cancer cases found in the state registry data. Most diagnosed cases (approx. 80%) were reported in either the follow-up surveys or the state registry data so we did not separate out national death index cases.

Next, we repeated our examination of the relationship between family history and colorectal cancer using a Cox proportional hazards model, a common choice for modeling time to medical events. For our analysis, the time to event was the diagnosis year minus the enrollment year for participants with a diagnosis of colorectal cancer and the censoring year minus the enrollment year for those without. The censoring year was the year of death (if applicable) or 2016, whichever occurred first.

Finally, we confirmed that our results were robust to altering the definition of family history. First, rather than grouping the participants with unknown family history with the “no family history” group, we grouped them with the “known family history” group. This might help address the mismeasurement of family history for Black participants if many of those with unknown family history did in fact have a family member with colorectal cancer. We also re-ran the analysis with family history as a categorical variable with three different categories: No, Don’t Know, and Yes.

All analyses were run in R version 4.2.1.

## RESULTS

Using data from the Southern Community Cohort Study (SCCS)^25^, established to study cancer disparities, we analyzed 77,836 adults (ages 40-74) with no history of colorectal cancer at baseline; Table 1 provides descriptive statistics. More than two-thirds of participants identified as Black/African-American and the rest as White. Approximately 7% of participants reported a known family history of colorectal cancer: 8.4% of White participants compared to 5.9% of Black participants. 7.2% of Black participants reported unknown family history, compared to 4.4% of White participants. The remaining participants reported no family history. 10-year colorectal cancer rates were higher in Black participants (1.9%) than White participants (1.6%). The fact that Black participants had higher colorectal cancer risk but *lower* rates of known colorectal cancer family history (and higher rates of unknown family history) suggests that family history information was less reliably recorded for Black participants.

**Table 1:** Sample Summary.

| **Variable** | **Black**<br>**Participants** | **White**<br>**Participants** | **All**<br>**Participants** |
| --- | --- | --- | --- |
| Female (%) | 58.4 | 61.1 | 59.3 |
| Enrollment Age (%) |  |  |  |
| 40-49 | 48.6 | 37.7 | 45.2 |
| 50-59 | 35.2 | 36.2 | 35.5 |
| 60-69 | 13.5 | 21.9 | 16.1 |
| 70-74 | 2.7 | 4.1 | 3.1 |
| Race (%) |  |  |  |
| Black | 100.0 | 0 | 69.1 |
| White | 0 | 100.0 | 30.9 |
| Family History of Colorectal Cancer (%) |  |  |  |
| Yes | 5.9 | 8.4 | 6.7 |
| Don't Know | 7.2 | 4.4 | 6.3 |
| Colorectal Cancer, 10-year (%) | 1.5 | 1.3 | 1.4 |
| Colorectal Cancer, Ever (%) | 2.0 | 1.6 | 1.9 |
| Mortality (%) | 25.3 | 27.1 | 25.9 |
| Number of Participants | 53,805 | 24,031 | 77,836 |

We compared the prognostic value of reported family history for 10-year colorectal cancer risk for Black versus White participants (Figure 1). Family history was strongly predictive of cancer risk for white participants (Logistic Regression odds ratio (OR): 1.74, 95% CI: 1.25-2.38, p-value 0.001) but not for Black participants (OR: 0.98, 95% CI: 0.72-1.29, p-value 0.887).

**Figure 1.**
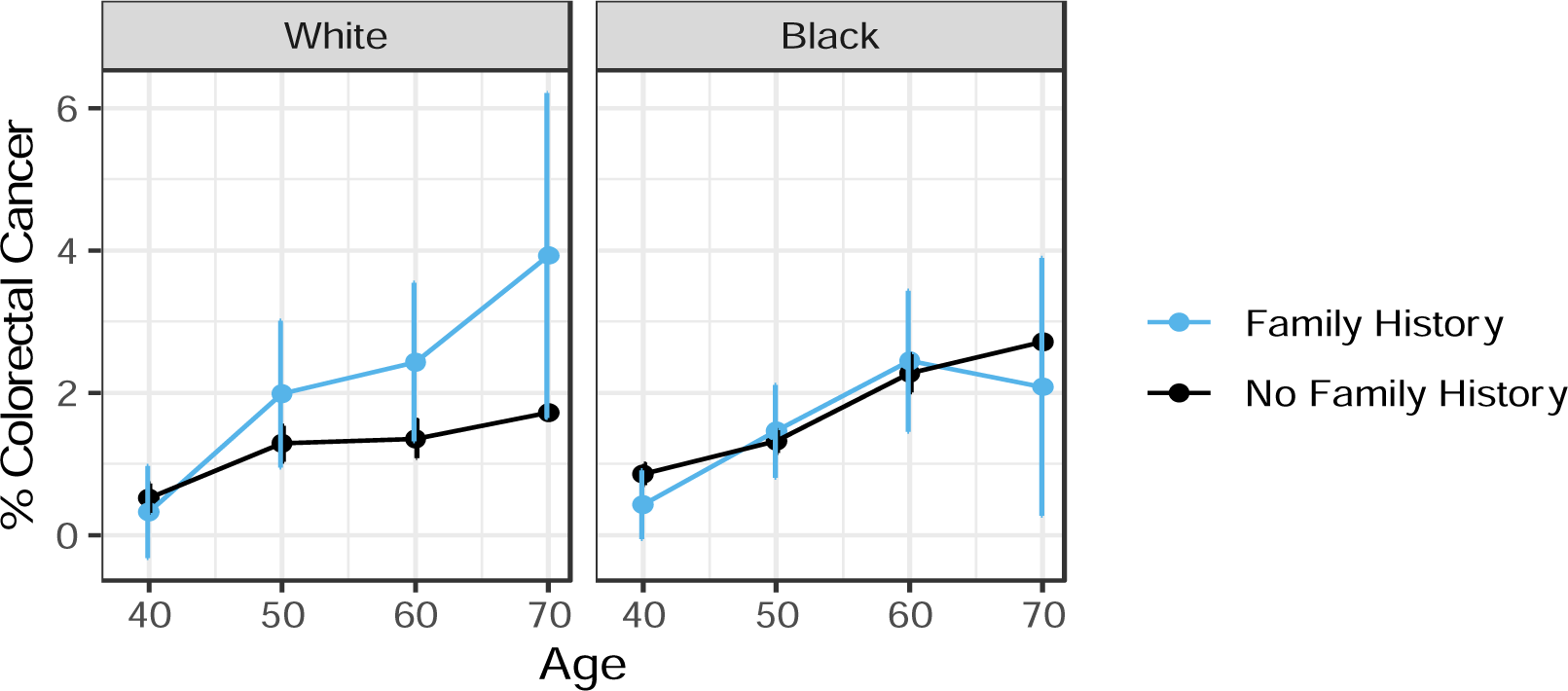
10-Year Colorectal Cancer Rates by Age, Family History, and Race. *Note:* Family history was predictive of cancer risk for White participants, but not Black participants.

Then we compared the race-blind algorithm to the race-corrected algorithm, which included race as a main effect and as an interaction with reported family history. In the race-corrected algorithm, both race terms were statistically significant: the main effect (p-value: <0.001) conveyed the fact that, among participants who did not report known family history, Black participants had 1.38x higher odds than White participants of developing colorectal cancer. The interaction term between race and family history was also statistically significant (p-value: 0.010), indicating that reported family history was more predictive in White patients than Black patients, consistent with what would be expected if family history were less reliably reported in Black participants. These results were consistent across alternative model specifications, outcome definitions, and family history definitions (Tables S1-S4).

The race-corrected algorithm improved several measures of prediction performance when compared to the race-blind algorithm. First, the race-corrected algorithm significantly improved goodness of fit (likelihood ratio test, p-value <0.001). Second, on a held-out test set, the race-corrected algorithm improved Area Under the Receiving Operating Characteristic (AUROC) among Black participants (0.611 versus 0.608, p-value: 0.006, DeLong’s method) and in the overall cohort (0.613 versus 0.606, p-value: 0.057), though the increase in AUROC among the overall cohort was only statistically significant at the p<0.10 level (Table 2). Finally, as illustrated in Figure S1, the race-blind algorithm underpredicted risks for Black patients, while the race-corrected algorithm was better-calibrated.

**Table 2.** AUROC in Race-Blind versus Race-Corrected Algorithm in Holdout Sample.

| **Participants** | **Race-Blind**<br>**AUC** | **Race-Corrected**<br>**AUC** | **Increase in**<br>**AUC** | **P Value for**<br>**2-sided test** |
| --- | --- | --- | --- | --- |
| All | 0.606 | 0.613 | 0.007 | 0.057 . |
| Black | 0.608 | 0.611 | 0.003 | 0.006** |
| White | 0.612 | 0.613 | 0.002 | 0.586 |
| P Value: <0.001 '***' <0.01. '**' <0.05 '*' < 0.1 '.' |  |  |  |  |

*Note:* P-values were calculated using the function *roc.test* in the R package pROC<sup>30</sup>, which compared paired ROC curves using DeLong's algorithm.

Previous work has raised concerns that race corrections could increase health disparities by, for example, moving Black patients to lower risk categories and thereby reducing access to screening or other preventive services.^1^ However, in our setting, we found the *opposite* effect: the race-corrected algorithm included a larger share of Black participants among the predicted high-risk group (Figure 2). With the race-corrected algorithm, 74.4% of participants flagged in the top 50% of predicted risk were Black compared to 66.1% with the race-blind algorithm (p-value: <0.001). Similar results held across all high-risk cutoffs (top quartile, top decile, and top percentile). This is consistent with the fact that the race-blind algorithm underpredicted risks for Black patients, while the race-corrected algorithm was better calibrated.

**Figure 2.**
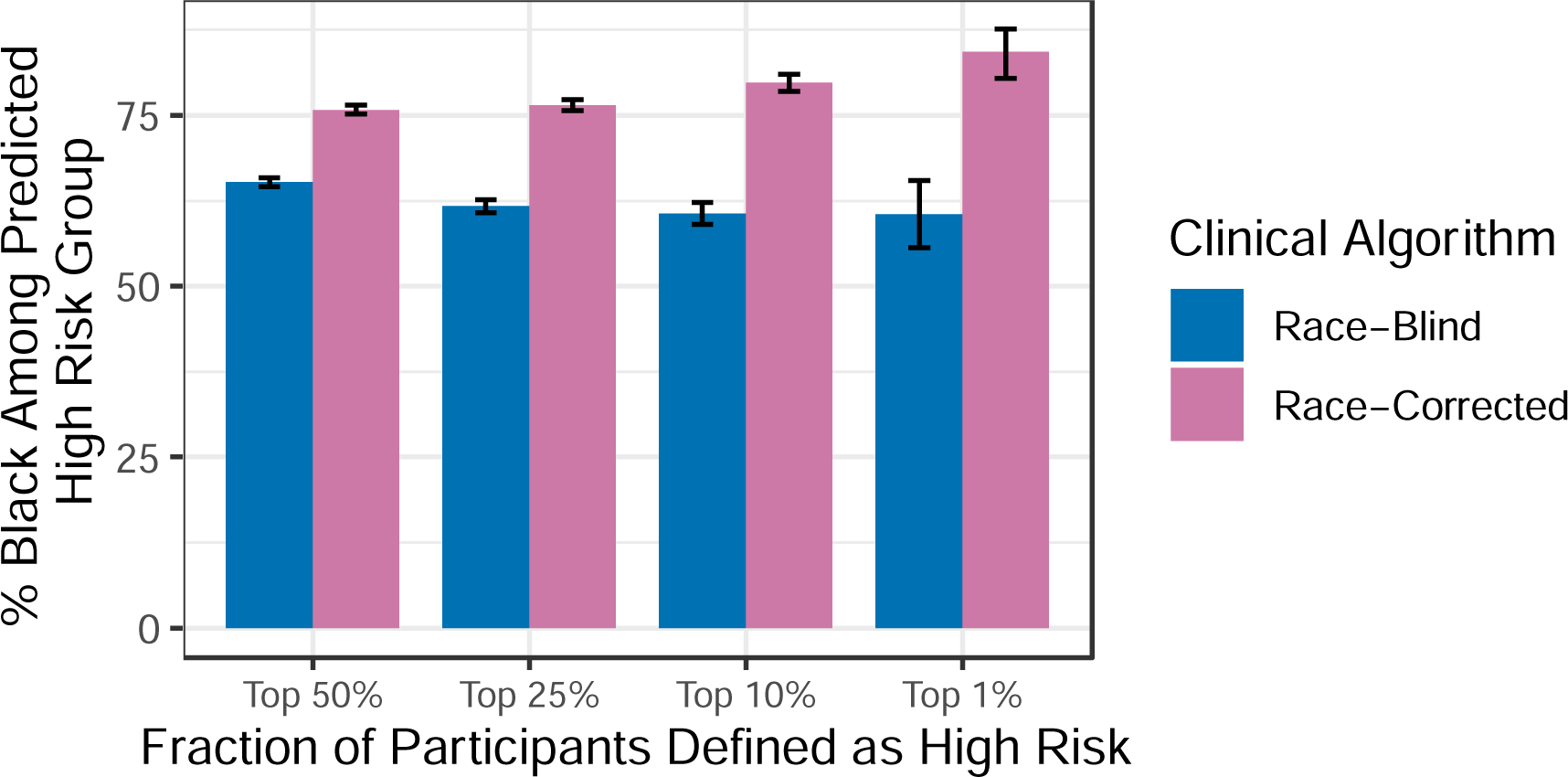
% Black Among Predicted High Risk Group, by High Risk Percentiles. *Note:* The race-corrected algorithm included more Black participants among the predicted high-risk group than the race-blind algorithm.

## DISCUSSION

Identifying individuals at high risk for colorectal cancer is an important component of prevention and screening practices in the U.S., where colorectal cancer remains the third leading cause of cancer-related death.^26^ In 2021, the United States Preventive Services Taskforce changed the recommended age for colorectal screening from 50 to 45 in the hopes of increasing screening rates to counteract early-onset cancer, among Black men in particular^27,28^, consistent with findings that colorectal cancer risk varies by race.^5^ Our analysis found that removing race from colorectal screening predictors could reduce the number of Black patients recommended for screening, which would work against efforts to reduce disparities in colorectal cancer screening and outcomes.

Overall, our analysis shows that including race in a colorectal cancer screening algorithm improved model fit, captured the differential predictive value of family history across groups, and increased the fraction of Black participants flagged as high risk, potentially increasing their access to screening. More broadly, when key inputs to clinical algorithms are mismeasured by race group, race correction can help correct for race disparities in the quality of health care data.

Our study has several limitations. First, although we showed an increase in the proportion of Black patients among the predicted high-risk group using a race-corrected algorithm, we do not assess how that would affect access to colorectal screenings for high-risk Black patients in real clinical settings, a direction for future work. Second, we analyze a specific clinical setting – colorectal cancer prediction – showing how race correction can improve clinical algorithms by accounting for data quality differences by race; future work should examine the same phenomenon in other clinical settings, given the pervasive and well-documented differences in clinical data quality by race group. Third, while we document a setting where race corrections improve predictive performance without increasing health disparities, in other settings, race corrections may not improve predictive performance or may perpetuate health disparities.^1^ Finally, while our study focuses on the use of race corrections to address differences in data quality by race group, other solutions such as better data collection should also be pursued in addition to, or in place of, race correction.^29^

In conclusion, our study illustrates how race corrections can help clinical algorithms capture varying data quality across race groups, a frequent phenomenon in health data. This argument does not rely on treating race as a biological variable, which it is not. Rather, our argument is about the effects of structural racism on the quality of medical data. The deep structural injustices which pervade healthcare and public health mean that critical inputs to medical algorithms -- like family history -- are more likely to be missing or misrecorded for some race groups. We do a disservice to patients if we fail to acknowledge this reality in our algorithms. But acknowledging that reality is not the same as accepting it. While race corrections can allow algorithms to capture current deficiencies in medical data, we need to look beyond that to what medical data could be, and must be, if we are to achieve health equity for all patients.

## Data Availability

Data are not available under the data use agreement.

## References

1. Vyas DA, Eisenstein LG, Jones DS. Hidden in Plain Sight — Reconsidering the Use of Race Correction in Clinical Algorithms. N Engl J Med. 2020;383(9):874–882. doi:10.1056/NEJMms2004740

2. Removing the Racial Coefficient From Calculations of eGFR. Accessed June 3, 2023. https://reports.mountsinai.org/article/neph2021-07-egfr-and-race

3. Diao JA, Wu GJ, Taylor HA, et al. Clinical Implications of Removing Race From Estimates of Kidney Function. JAMA. 2021;325(2):184–186. doi:10.1001/jama.2020.22124

4. Embracing Genetic Diversity to Improve Black Health | NEJM. Accessed June 3, 2023. https://www.nejm.org/doi/full/10.1056/NEJMms2031080

5. Khor S, Haupt EC, Hahn EE, Lyons LJL, Shankaran V, Bansal A. Racial and Ethnic Bias in Risk Prediction Models for Colorectal Cancer Recurrence When Race and Ethnicity Are Omitted as Predictors. JAMA Netw Open. 2023;6(6):e2318495. doi:10.1001/jamanetworkopen.2023.18495

6. Bonner SN, Lagisetty K, Reddy RM, Engeda Y, Griggs JJ, Valley TS. Clinical Implications of Removing Race-Corrected Pulmonary Function Tests for African American Patients Requiring Surgery for Lung Cancer. JAMA Surg. Published online August 16, 2023:e233239. doi:10.1001/jamasurg.2023.3239

7. Chen IY, Pierson E, Rose S, Joshi S, Ferryman K, Ghassemi M. Ethical Machine Learning in Healthcare. Annu Rev Biomed Data Sci. 2021;4(1):null. doi:10.1146/annurev-biodatasci-092820-114757

8. Greenland S. The effect of misclassification in the presence of covariates. Am J Epidemiol. 1980;112(4):564–569. doi:10.1093/oxfordjournals.aje.a113025

9. Clarke R, Shipley M, Lewington S, et al. Underestimation of risk associations due to regression dilution in long-term follow-up of prospective studies. Am J Epidemiol. 1999;150(4):341–353. doi:10.1093/oxfordjournals.aje.a010013

10. Hutcheon JA, Chiolero A, Hanley JA. Random measurement error and regression dilution bias. BMJ. 2010;340:c2289. doi:10.1136/bmj.c2289

11. Family Health History and Cancer. Centers for Disease Control and Prevention. Published January 9, 2023. Accessed May 6, 2023. https://www.cdc.gov/cancer/family-health-history/index.htm

12. Murff HJ, Byrne D, Haas JS, Puopolo AL, Brennan TA. Race and Family History Assessment for Breast Cancer. J Gen Intern Med. 2005;20(1):75–80. doi:10.1111/j.1525-1497.2004.40112.x

13. Kupfer SS, McCaffrey S, Kim KE. Racial and gender disparities in hereditary colorectal cancer risk assessment: the role of family history. J Cancer Educ Off J Am Assoc Cancer Educ. 2006;21(1 Suppl):S32-36. doi:10.1207/s15430154jce2101s_7

14. Chavez-Yenter D, Goodman MS, Chen Y, et al. Association of Disparities in Family History and Family Cancer History in the Electronic Health Record With Sex, Race, Hispanic or Latino Ethnicity, and Language Preference in 2 Large US Health Care Systems. JAMA Netw Open. 2022;5(10):e2234574. doi:10.1001/jamanetworkopen.2022.34574

15. Andoh JE. The Stories We Don’t Know. JAMA. Published online April 13, 2023. doi:10.1001/jama.2023.5891

16. Colorectal Cancer Guideline | How Often to Have Screening Tests. Accessed May 11, 2022. https://www.cancer.org/cancer/colon-rectal-cancer/detection-diagnosis-staging/acs-recommendations.html

17. Southern Community Cohort Study. Southern Community Cohort Study. Published 2022. Accessed November 1, 2022. https://www.southerncommunitystudy.org/

18. Win AK, MacInnis RJ, Hopper JL, Jenkins MA. Risk Prediction Models for Colorectal Cancer: A Review. Cancer Epidemiol Biomarkers Prev. 2012;21(3):398–410. doi:10.1158/1055-9965.EPI-11-0771

19. Obermeyer Z, Powers B, Vogeli C, Mullainathan S. Dissecting racial bias in an algorithm used to manage the health of populations. Science. 2019;366(6464):447-453. doi:10.1126/science.aax2342

20. Amersi F, Agustin M, Ko CY. Colorectal Cancer: Epidemiology, Risk Factors, and Health Services. Clin Colon Rectal Surg. 2005;18(3):133–140. doi:10.1055/s-2005-916274

21. Colorectal Cancer Risk Assessment Tool. Colorectal Cancer Risk Assessment Tool. Published 2022. Accessed May 13, 2022. https://www.cancer.gov/ccrisktool

22. Hajian-Tilaki K. Receiver Operating Characteristic (ROC) Curve Analysis for Medical Diagnostic Test Evaluation. Casp J Intern Med. 2013;4(2):627–635.

23. DeLong ER, DeLong DM, Clarke-Pearson DL. Comparing the Areas under Two or More Correlated Receiver Operating Characteristic Curves: A Nonparametric Approach. Biometrics. 1988;44(3):837–845. doi:10.2307/2531595

24. Abd ElHafeez S, D’Arrigo G, Leonardis D, Fusaro M, Tripepi G, Roumeliotis S. Methods to Analyze Time-to-Event Data: The Cox Regression Analysis. Oxid Med Cell Longev. 2021;2021:e1302811. doi:10.1155/2021/1302811

25. Signorello LB, Hargreaves MK, Blot WJ. The Southern Community Cohort Study: investigating health disparities. J Health Care Poor Underserved. 2010;21(1 Suppl):26-37. doi:10.1353/hpu.0.0245

26. American Cancer Society. Colorectal Cancer Statistics | How Common Is Colorectal Cancer? Accessed May 11, 2022. https://www.cancer.org/cancer/colon-rectal-cancer/about/key-statistics.html

27. Mauri G, Sartore-Bianchi A, Russo AG, Marsoni S, Bardelli A, Siena S. Early-onset colorectal cancer in young individuals. Mol Oncol. 2019;13(2):109–131. doi:10.1002/1878-0261.12417

28. MPH AC MD. Racial disparities and early-onset colorectal cancer: A call to action. Harvard Health. Published March 17, 2021. Accessed May 11, 2022. https://www.health.harvard.edu/blog/racial-disparities-and-early-onset-colorectal-cancer-a-call-to-action-202103172411

29. Berglund L. Regression dilution bias: Tools for correction methods and sample size calculation. Ups J Med Sci. 2012;117(3):279–283. doi:10.3109/03009734.2012.668143

30. Robin X, Turck N, Hainard A, et al. pROC: an open-source package for R and S+ to analyze and compare ROC curves. BMC Bioinformatics. 2011;12(1):77. doi:10.1186/1471-2105-12-77

