## Supplement for "Race Corrections in Clinical Algorithms Can Help Correct for Racial Disparities in Data Quality"

for

“**Race Corrections in Clinical Algorithms**

**Table S1. Odds Ratio (95% Confidence Intervals) for Logistic Regression Predicting 10-Year Colorectal Cancer**

| **Variables** | **(1)**  **Black Participants** | **(2)**  **White**  **Participants** | **(3)**  **Race-Blind Algorithm** | **(4)**  **Race-Corrected Algorithm** |
| --- | --- | --- | --- | --- |
| *All Data* | |  |  |  |
| Family History | 0.979  (0.724 to 1.293) | 1.743**  (1.246 to 2.383) | 1.255*  (1.006 to 1.548) | 1.802***  (1.285 to 2.467) |
| Black |  |  |  | 1.376***  (1.192 to 1.593) |
| Family History × Black**^†^** |  |  |  | 0.564*  (0.366 to 0.873) |
| *Follow-Up Survey Data***^‡^** | |  |  |  |
| Family History | 0.939  (0.604 to 1.389) | 2.068**  (1.376 to 3.007) | 1.317 .  (0.983 to 1.729) | 2.022***   (1.343 to 2.949) |
| Black |  |  |  | 1.205 .  (0.997 to 1.463) |
| Family History × Black**^†^** |  |  |  | 0.458**  (0.257 to 0.807) |
| *Cancer Registry Data* | |  |  |  |
| Family History | 1.068  (0.751 to 1.473) | 1.626*  (1.065 to 2.390) | 1.328*  (1.017 to 1.705) | 1.794**  (1.173 to 2.643) |
| Black |  |  |  | 1.370***  (1.151 to 1.638) |
| Family History × Black**^†^** |  |  |  | 0.633  (0.375 to 1.077) |
| Age Controls | Y | Y | Y | Y |
| Full NIH Controls |  |  | Y | Y |

P Value: <0.001 ‘***’	<0.01 ‘**’	<0.05 ‘*’	< 0.1 ‘.’

**^†^**Family History × Black is the coefficient on the interaction between family history and an indicator for Black race.

**^‡^**Excludes 31.7% of the sample that didn’t complete follow-up surveys.

*Note:* The coefficient on the interaction term remains similar across all three specifications (though the confidence interval is wider in the final specification and thus not statistically significant) indicating that, across our robustness checks, family history was consistently less predictive for Black than White patients.

**Table S2. Hazard Ratios (95% Confidence Intervals) for Cox Proportional Hazard Model Predicting 10-Year Colorectal Cancer**

| **Variables** | **(1)**  **Black Participants** | **(2)**  **White**  **Participants** | **(3)**  **Race-Blind Algorithm** | **(4)**  **Race-Corrected Algorithm** |
| --- | --- | --- | --- | --- |
| Family History | 0.995  (0.765 to 1.296) | 1.722***  (1.275 to 2.326) | 1.254*  (1.029 to 1.528) | 1.766***   (1.306 to 2.387) |
| Black |  |  |  | 1.304***  (1.139 to 1.493) |
| Family History × Black**^†^** |  |  |  | 0.583**   (0.391 to 0.870) |
| Age Controls | Y | Y | Y | Y |
| Full NIH Controls |  |  | Y | Y |

P Value: <0.001 ‘***’	<0.01 ‘**’	<0.05 ‘*’	< 0.1 ‘.’

**^†^**Family History × Black is the coefficient on the interaction between family history and an indicator for Black race.

*Note:* The coefficient on the interaction term remains similar using a cox proportional hazard model indicating that, across our robustness checks, family history was consistently less predictive for Black than White patients.

**Table S3. Odds Ratio (95% Confidence Intervals) for Logistic Regression Predicting 10-Year Colorectal Cancer using Alternative Family History Definition**

| **Variables** | **(1)**  **Black Participants** | **(2)**  **White**  **Participants** | **(4)**  **Race-Blind Algorithm** | **(5)**  **Race-Corrected Algorithm** |
| --- | --- | --- | --- | --- |
| Family History**^†^** | 1.100  (0.901 to 1.331) | 1.596**   (1.189 to 2.108) | 1.237*  (1.050 to 1.449) | 1.591**  (1.185 to 2.103) |
| Black |  |  |  | 1.372***  (1.183 to 1.597) |
| Family History × Black**^†^** |  |  |  | 0.695*   (0.494 to 0.987) |
| Age Controls | Y | Y | Y | Y |
| Full NIH Controls |  |  | Y | Y |

P Value: <0.001 ‘***’	<0.01 ‘**’	<0.05 ‘*’	< 0.1 ‘.’

**^†^**Family history used here groups participants who don’t know whether a family member has had colorectal cancer or not with participants who report a known family history. Family History × Black is the coefficient on the interaction between family history and an indicator for Black race.

*Note:* The coefficient on the interaction term remains similar when we use an alternative definition of family history indicating that, across our robustness checks, family history was consistently less predictive for Black than White patients.

**Table S4. Odds Ratio (95% Confidence Intervals) for Logistic Regression Predicting 10-Year Colorectal Cancer using 3-level Categorical Family History Definition**

| **Variables** | **(1)**  **Black Participants** | **(2)**  **White Participants** | **(3)**  **Race-Blind**  **Algorithm** | **(4)**  **Race-Corrected Algorithm** |
| --- | --- | --- | --- | --- |
| Family History (Don’t Know) **^†^** | 1.187  (0.921 to 1.506) | 1.263  (0.728 to 2.033) | 1.201  (0.955 to 1.490) | 1.184  (0.682 to 1.190) |
| Family History (Yes) **^†^** | 0.995  (0.735 to 1.315) | 1.766**   (1.260 to 2.419) | 1.273*  (1.019 to 1.571) | 1.818***  (1.295 to 2.494) |
| Black |  |  |  | 1.371***  (1.182 to 1.595) |
| Family History (Don’t Know) × Black**^†^** |  |  |  | 0.985  (0.573 to 1.792) |
| Family History (Yes) × Black**^†^** |  |  |  | 0.566*  (0.366 to 0.878) |
| Age Controls | Y | Y | Y | Y |
| Full NIH Controls |  |  | Y | Y |

P Value: <0.001 ‘***’	<0.01 ‘**’	<0.05 ‘*’	< 0.1 ‘.’

**^†^**Family history used here has three categories (No, Don’t Know, and Yes). No family history is the reference group. Family History × Black is the coefficient on the interaction between the family history category and an indicator for Black race.

*Note:* The coefficient on the interaction term Family History (Yes) × Black remains similar when we use an alternative definition of family history indicating that, across our robustness checks, family history was consistently less predictive for Black than White patients.

**Figure S1. Actual vs. Predicted 10-Year Colorectal Cancer Risk**

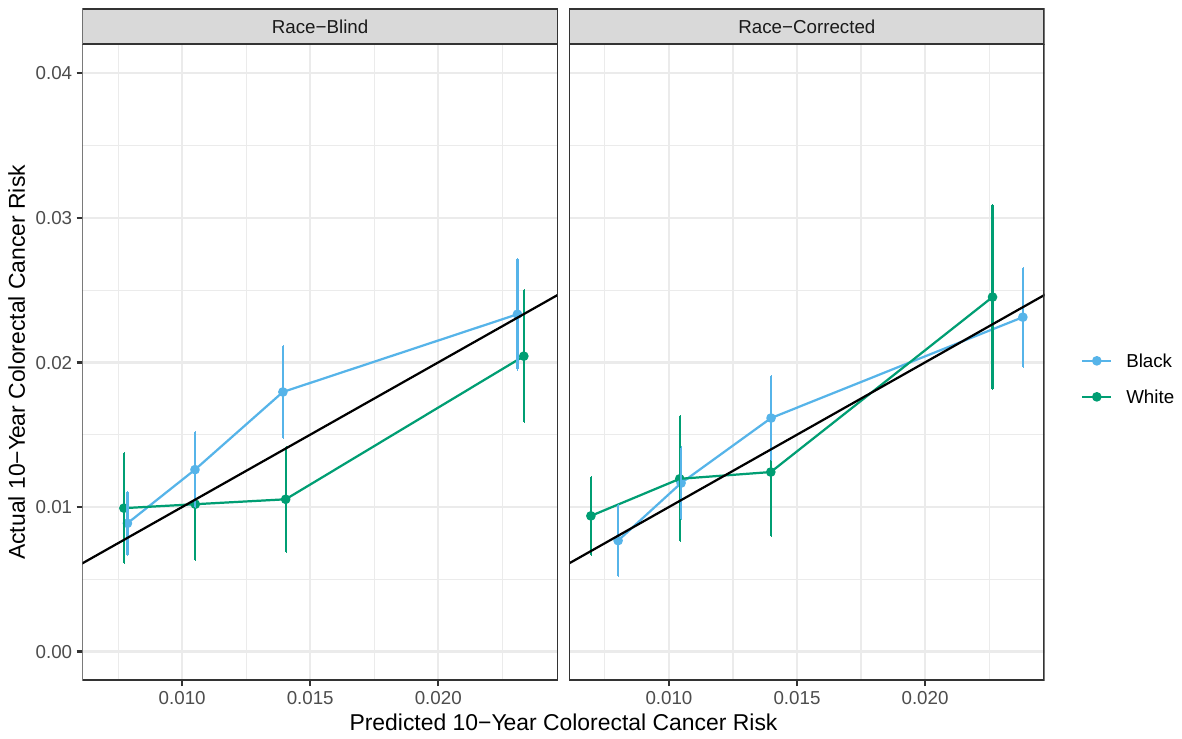

*Note:* The race-corrected algorithm (right) yielded better-calibrated estimates than the race-blind algorithm (left). The horizontal axis plots predicted risk, and the vertical axis plots true risk, for quartiles of the test set. Perfectly calibrated estimates, where predicted risk and true risk are equivalent, would lie along the black diagonal line. The race-blind algorithm yielded predictions which are too low for Black patients; in contrast, the race-corrected algorithm mitigated this issue (decrease in expected calibration error for Black patients: 0.001211, 95% CI: 0.0012-0.0028).
